# Effectiveness of the Ticket to Work program in supporting employment among adults with disabilities

**DOI:** 10.1101/2025.04.22.25325884

**Authors:** Pei-Shu Ho, Joshua C. Chang, Rebecca A. Parks, Kathleen Coale, Chunxiao Zhou, Rafael Jiménez Silva, Julia Porcino, Elizabeth Marfeo, Elizabeth K. Rasch

## Abstract

**Background:** Returning to work benefits many people with disabilities, as it supports personal financial independence and provides opportunities for greater societal contributions. The U.S. Social Security Administration’s Ticket to Work (TTW) program offers expanded support services to help disability beneficiaries achieve financial independence through gainful employment. SSA has continuously sought new ways to identify those who would most benefit from using a Ticket.

**Objective:** To identify factors contributing to TTW participation and assess its impact on benefits forgone for work.

**Methods:** We conducted a cohort study using SSA administrative data to predict TTW participation and its impact on benefit cessation. The study sample included beneficiaries with a physical or mental residual functional capacity assessment from 2016. We applied a frequentist propensity score matching estimate and a doubly robust Bayesian hierarchical model-based estimate.

**Results:** The study included 172,640 beneficiaries (52.7% male, average age 52 years) with a range of qualifying conditions: musculoskeletal disorders (45.04%), mental disorders (29.10%), neurological disorders (9.82%). Both analytic methods yielded consistent results, showing that TTW participation is effective even after controlling for confounding factors. Personal characteristics (e.g., sex, age, education, race/ethnicity), health and functional status (e.g., work cessation due to health issues, need for alternate sitting arrangements, and limitations in understanding and memory) and environmental factors (e.g., region of residence) influenced Ticket participation.

**Conclusions:** Our findings offer valuable insights for identifying potential TTW participants and estimating benefit savings for SSDI/SSI programs. Future research should explore available support services and barriers to access to improve TTW outcomes.

## Introduction

Employment is a key source of personal identity, financial stability, and social connection.^1^ It offers numerous benefits to individuals and society, including self-sufficiency, social interaction, improved quality of life, and contributions to the community through goods and services. However, individuals with disabilities are less likely to work compared to those without disabilities. According to a recent news release from the Bureau of Labor Statistics, only about 37% of working age adults (ages 16-64) with disabilities worked in 2023 - roughly half the employment rate of their counterparts without disabilities (75%).^2^ While many interventions to facilitate return-to-work (RTW) following an illness or accident have been studied, most of them have focused on individuals with specific health-related conditions or diagnoses, such as cancers^3,4^ stroke^5^, coronary heart disease,^6^ trauma,^7,8^ mental health,^7,9–12^ and musculoskeletal conditions.^12^ Factors influencing RTW also vary by condition. For example, among cancer survivors, lower fatigue levels, higher work value, stronger job self-efficacy, and greater perceived work ability are associated with earlier RTW.^13^ Among stroke survivors, RTW is associated with factors such as functional disability, job type,^14^ and fatigue level.^15^ Additional characteristics associated with the likelihood of RTW among individuals with disabilities include age, gender, race, education, use of assistive devices, physical functioning, severity of pain or symptoms, living situation, job demands and accommodations, access to health benefits, opportunities for advancement, and supervisors’ attitudes.^16–19^

To support the workforce re-entry of disability program beneficiaries, the Ticket to Work and Work Incentives Improvement Act (TTWIIA) of 1999 authorized the U.S. Social Security Administration (SSA) to implement the Ticket to Work and Self-Sufficiency (TTW) program.^20^This program targets working-age beneficiaries (ages 18-64) who receive Social Security Disability Insurance (SSDI) and/or Supplemental Security Income (SSI) disability benefits. TTW aims to (1) expand service and support options to facilitate employment; (2) promote financial independence and self-sufficiency through work; and (3) reduce reliance on disability benefits.^21^ Eligible beneficiaries may use a “Ticket” to access vocational rehabilitation, employment services from participating providers, including Employment Networks (ENs) and State Vocational Rehabilitation Agencies (SVRAs), which have formal agreements with SSA to deliver or coordinate these supports.

Since its launch in 2002, SSA has introduced several measures to enhance program participation. These include mailing Tickets directly to eligible beneficiaries, establishing a toll-free helpline, and waiving medical continuing disability reviews while the Ticket is in use.^22^ Additionally, SSA revised TTW regulations in 2008 to encourage provider participation by offering larger payments for lower-level earnings, earlier payments in the RTW process, and streamlined administrative requirements for ENs.^23^ These changes led to modest increases in participation by both beneficiaries and service providers. Despite these efforts, overall TTW participation remains limited, and the program’s impact on employment outcomes remains mixed.^24–31^ Previous evaluations have called for better use of multiple sources of SSA administrative data to identify the characteristics of beneficiaries most likely to participate in TTW and to benefit from it.^22^

Therefore, the present study aims to analyze SSA’s administrative data to (1) identify characteristics of beneficiaries most likely to participate in the TTW program and (2) examine how beneficiaries’ characteristics and TTW participation are associated with the likelihood of RTW. Findings from this research aim to inform strategies for improving employment outcomes among working-age adults with disabilities and optimizing the effectiveness of employment support programs like TTW.

## Methods

### Data source

This study used data from the SSA 2021 Disability Analysis File (DAF), the electronic Claims Analysis Tool (eCAT), and the electronic Disability System (eDib). The DAF^32^ includes longitudinal and one-time administrative data on SSDI and SSI beneficiaries, such as program participation and benefit status. We used several DAF components, including Awardee Data Mart, the Demographic File, Annual Files, and Ticket to Work Beneficiary Participation Files. Key measures, such as monthly indicators for program participation, suspension or termination for work, and benefits foregone for work are pre-constructed within the dataset. The eCAT provides assessments of Physical and Mental Residual Functional Capacity (RFC). Physical RFC (PRFC) covers six domains: exertional, postural, manipulative, visual, communicative, and environmental limitations. The Mental RFC (MRFC) evaluates four domains: understanding and memory, sustained concentration and persistence, social interaction, and adaptation limitations. The eDib supports SSA’s claims processing^33^ and includes the SSA-3368-BK and SSA-3369-BK forms, which provide self-reported information on health, education and training history, work activity, and employment history.

### Study population

We included beneficiaries allowed at Step 5 of the SSA’s sequential disability determination process in 2016, followed through 2021. Beneficiaries allowed at Step 5 are based on a person’s inability to perform any job in the national economy, factoring in age, education, work experience, and residual functional capacity.^34^ After excluding deceased individuals, the total final analytic sample included 172,640 beneficiaries (see Supplementary Materials).

### Conceptual framework

Our analysis was guided by the Illinois Work and Well-Being Model,^35^ which views workforce participation as shaped by personal, environmental, and systemic factors. The model highlights how contextual characteristics (e.g., age, education, functional capacity), career development factors (e.g., job seeking and job maintaining skills), and potential interventions (e.g., external supports), influence overall societal participation (including work or community participation). Based on this framework, we hypothesized that beneficiaries’ socio-demographic characteristics, health and functional status, environmental context, TTW participation, awareness of TTW program, employment history, and access to vocational services are associated with workforce participation.

### Measures

We constructed a binary dependent variable of TTW participation – that is, whether they received services from SVRAs or ENs. The independent variables were grouped into three categories: personal factors, health and functional limitations, and environmental factors. Personal factors included age at earliest benefit award, sex (male, female), race and ethnicity (Hispanic White, Hispanic Black, Hispanic, and other races), education (less than high school, high school, above high school), type of occupation, and work experience. Health and functional limitations comprised primary diagnosis, body mass index (BMI), any instance of overnight hospital stay, presence of pain, duration on benefit rolls, and Physical and Mental RFC measures. Environmental factors consisted of living arrangement (living in own house, alone, or having no dependents) and region of residence (Northeast, Midwest, South, West, U.S Territories).

Based on work history, we classified employment status into three categories: always worked full-time, always worked part-time, or worked both full-time and part-time. To standardize self-reported occupations, we applied the National Institute for Occupational Safety and Health (NIOSH) Industry and 2018 Occupation Computerized Coding System (NIOCCS), which converts job titles into 23 standard occupation codes. We selected the five most common occupations for analysis. Similarly, we grouped primary impairment codes by body system and analyzed the five most prevalent categories.

Using self-reported weight and height, we classified BMI into underweight (<18.5), healthy weight (18.5 to <25), overweight (25 to <30), class 1 obesity (30 to <35), class 2 obesity (35 to <40), and class 3 obesity (≥40).

As previously noted, the PRFC assessment evaluates six areas of physical functioning. Each category includes up to eight items on scales reflecting weight, frequency, or duration. For the analysis, we combined certain items into composite measures. Specifically, exertional limitations were summarized into four variables: (1) lifting/carrying objects; (2) standing/walking (with or without assistive devices); (3) sitting (with or without alternate positions); and (4) pushing/pulling. For the remaining categories - posture, manipulation, vision, communication, and environmental - we created summary measures representing the total number of limitations within each category.

The MRFC assessment covers four domains of mental functioning and includes 20 items rated on a four-point Likert scale: no, mild, moderate, and marked limitation. For each domain, we created a binary indicator denoting whether the beneficiary had a marked (i.e., serious) limitation.

We created a second binary dependent variable to capture whether any monthly benefits were forgone due to work between January 1, 2016 and December 31, 2021. A value of 1 indicated some benefits were forgone, while 0 indicated no suspension due to work. In this model, TTW participation was the independent variable, while the personal, health and functional, and environmental factors – previously used as independent variables – were included as covariates.

### Statistical analysis

We conducted descriptive analyses – including frequencies, percentages, means, standard deviations, and medians – to summarize beneficiaries’ characteristics. While we were interested in the predictive properties of each of our dependent variables, our primary focus was on estimating the causal inference of the effect of TTW participation on benefits suspension due to work. Because random assignment was not feasible with administrative data, we employed two methods to reduce confounding bias: frequentist propensity score matching (PSM) and Bayesian modeling. Comparing results across these approaches allowed us to assess the robustness of the findings.

Apart from the RFC variables, all other predictors for each of the fitted models were consistently specified across models. Some variables (e.g., age at first award) were recoded into binary categories, while others (e.g., English proficiency, residence in U.S. Territories) were excluded due to the small sizes or number of observations or limited variation.

#### Frequentist propensity score matching (PSM) analysis

To estimate the impact of TTW participation on benefits forgone due to work while reducing confounding bias, we applied the PSM within subgroups defined by SSI/SSDI status and RFC availability. For each subgroup, we first estimated the probability of TTW participation using a logistic regression model that included personal, health and functional, and environmental characteristics. These estimated probabilities (i.e., propensity scores) were then used for participant matching. We applied nearest neighbor matching with a 0.01 caliper on the logit of the propensity score to ensure high-quality matches and minimize residual confounding. Covariate balance was assessed before and after matching using standardized mean differences. Unmatched units were excluded from analysis. Using the matched samples, we estimated logistic regression models predicting benefit cessation due to work and computed adjusted estimates of the average treatment effect on the treated (ATT) of TTW participation. All these analyses were conducted using Stata.

#### Bayesian doubly robust causal inference

We also developed a Bayesian multilevel logistic regression model to control for confounding bias in estimating the effect of TTW participation on the outcome. This method differs from the frequentist approach in several ways. First, the Bayesian model encompasses all study participants simultaneously to avoid double-accounting for people with concurrent RFC data and to increase statistical power. Second, heterogeneity of the model parameters is inherently built into the model, allowing the different portions of the model to borrow information from each other. ^36,37^ Third, the Bayesian model is additionally regularized in a manner to promote sparsity by the incorporation of regularized horseshoe priors on the regression parameters.^38,39^ Fourth, the Bayesian model treats TTW participation (propensity score) and benefits suspension due to work as two outcomes that are modeled simultaneously – this model is a joint outcome analysis.^40,41^ Fifth, rather than using matching, confounding adjustment is performed directly within the model by incorporating a *clever covariate*^42^ as a predictor in benefit cessation.

The *clever covariate* is a function of both the propensity score and the treatment indicator. When using it as a predictor in outcome regression one obtains a doubly robust estimate of the average treatment effect in the sense that only one of the outcome or treatment models needs to be well-specified. One can then compute the overall average treatment effect for any cohort of individuals modeled by summing a contribution due to the clever covariate and a contribution due to the treatment variable.^43^

For Bayesian inference, we used a combination of the BlackJax^44^ Python package and Tensorflow Probability,^45^ with the aid of the bayesianquilts^37^ python package. To evaluate model accuracy in the context of generalizability, we report leave-on-out cross-validated area on the receiver curve (AUROC) metrics computed using importance sampling.^46^

## Results

### Characteristics of beneficiaries

The average age of the 172,640 beneficiaries at their earliest award was approximately 52 years (SD=12.92) (Table 1). Over half were males (52.71%), non-Hispanic White (60.09%), high school graduates (66.24%), born in the United States (87.90%), and living alone or in their own house (74.64%). More than one-third of beneficiaries resided in the South (37.03%). The three most common primary diagnostic categories were musculoskeletal disorders (45.04%), mental disorders (29.10%), and neurological disorders (9.82%). A substantial proportion of beneficiaries was obese (i.e., BMI≥30, 44.09%) and experienced pain or other symptoms related to their conditions (91.85%). Additionally, nearly 70 percent reported stopping work due to poor health (69.46%).

**Table 1.**
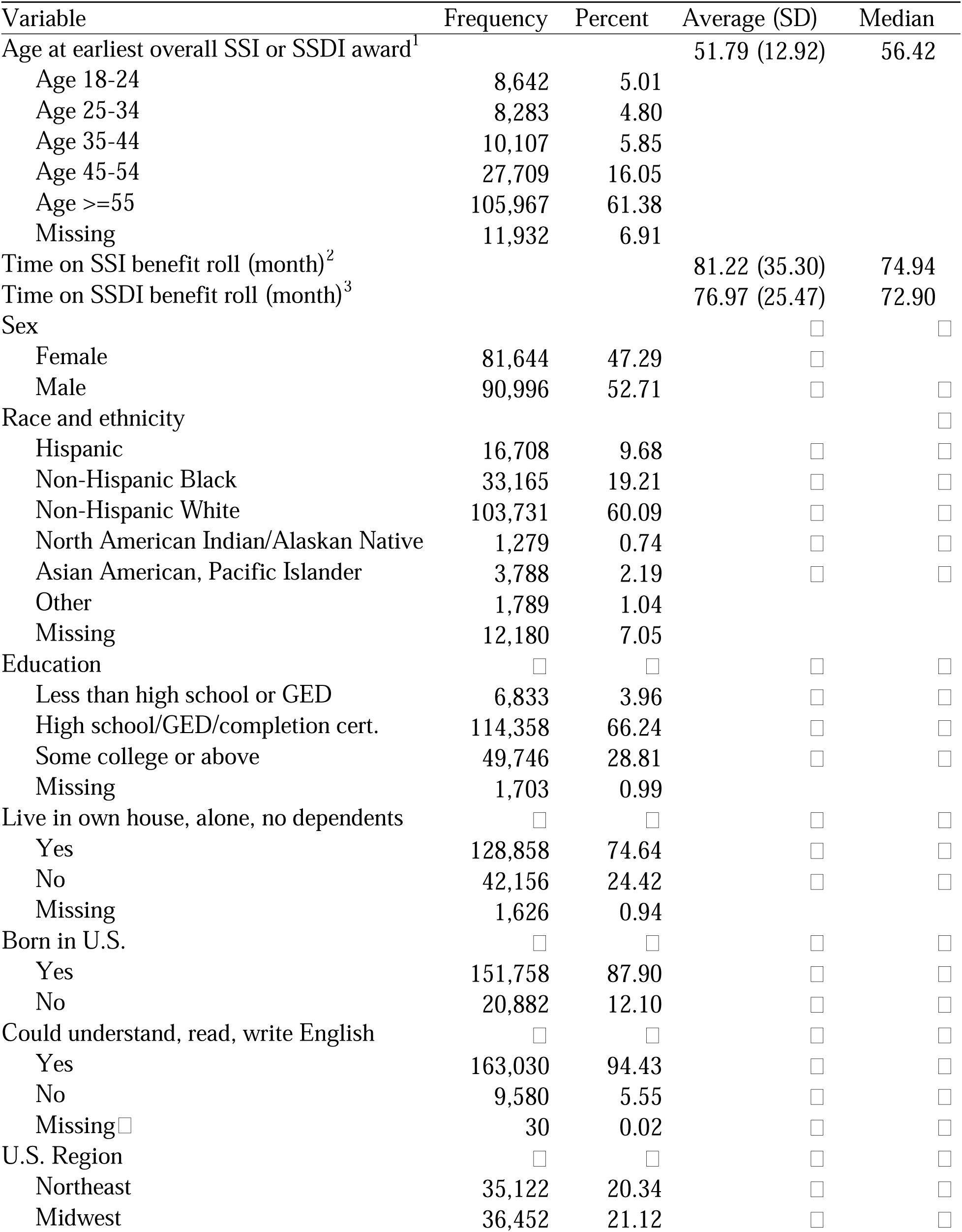
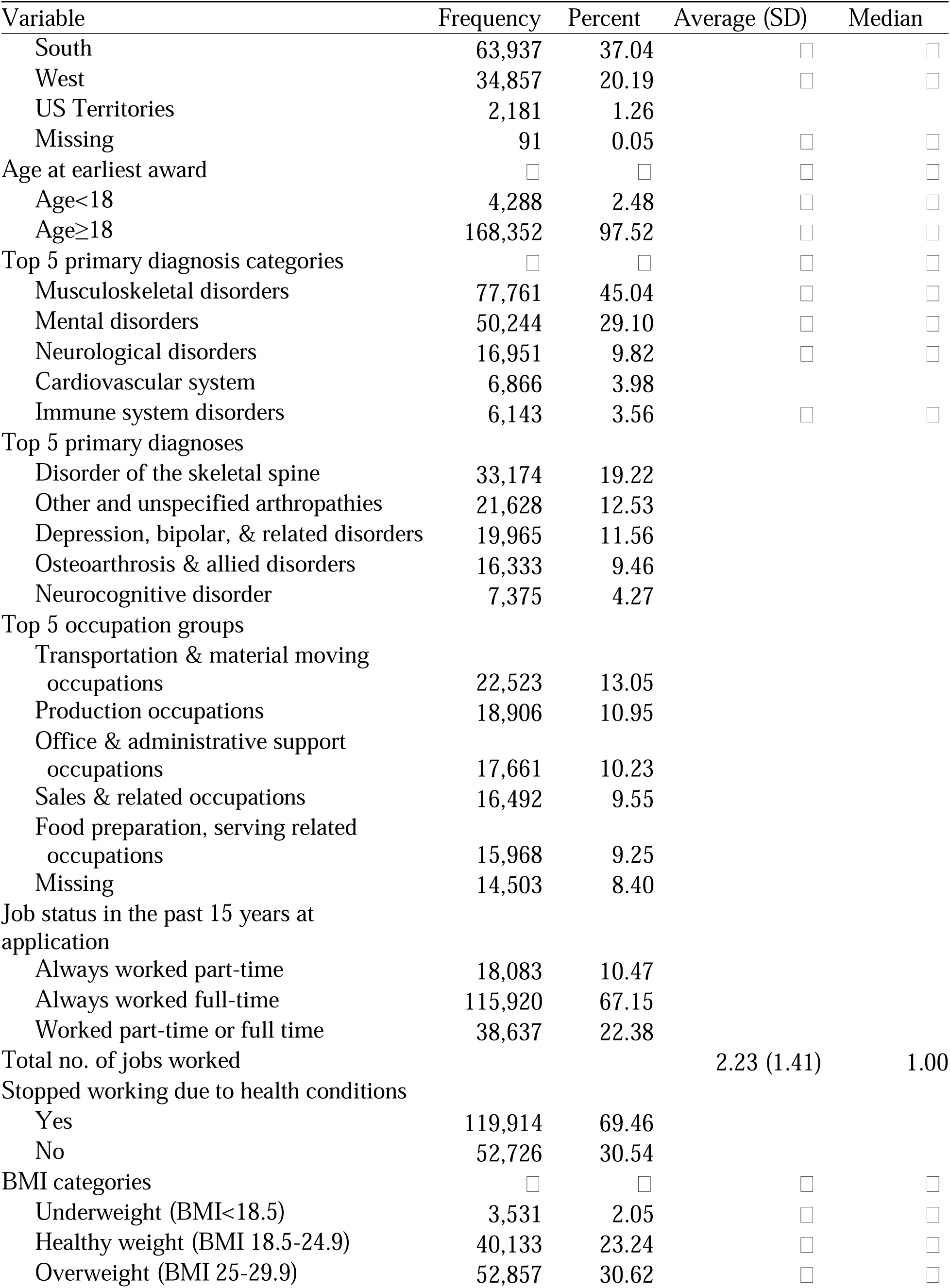
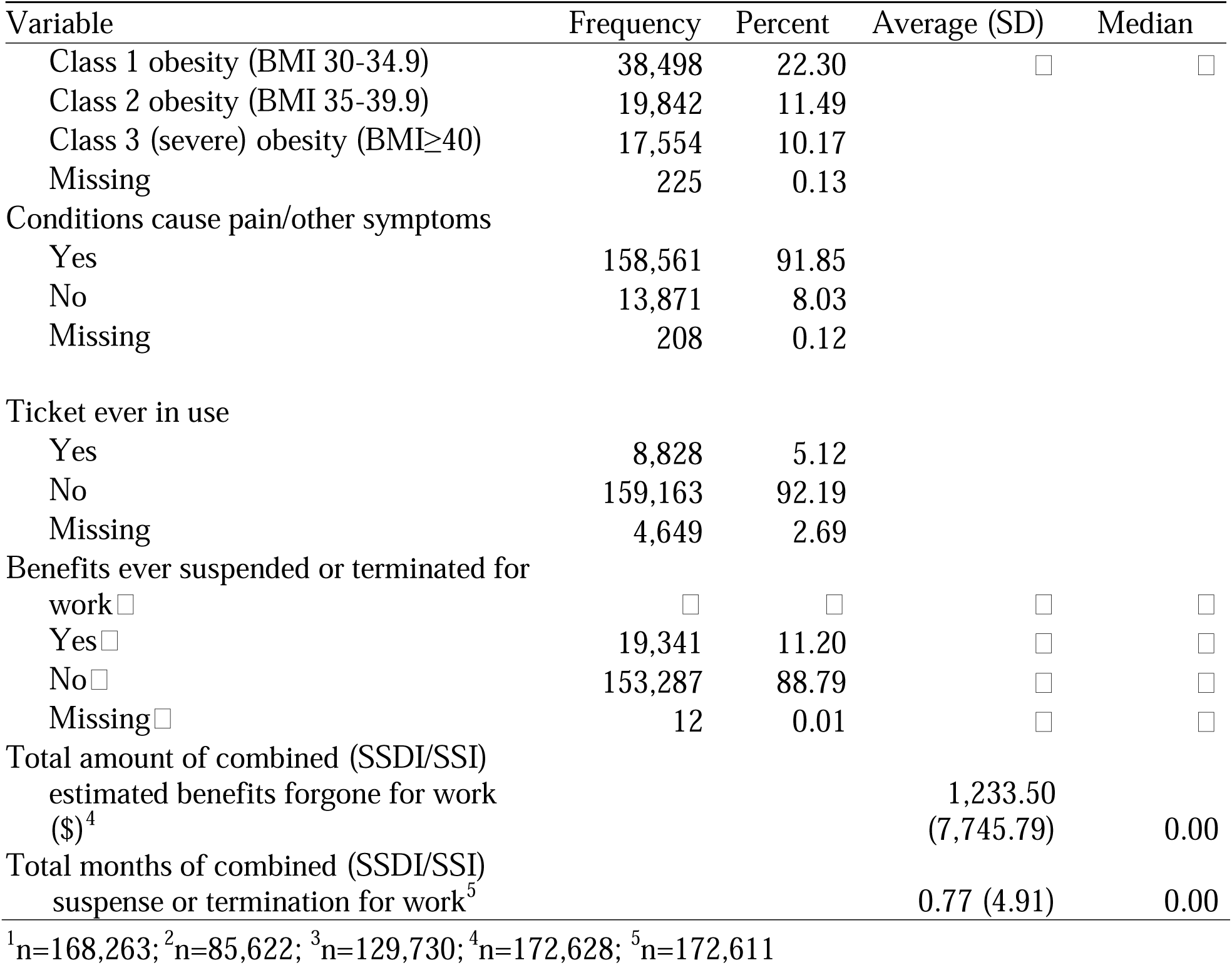
Characteristics of Study Sample (n=172,640)

| Variable | Frequency | Percent | Average (SD) | Median |
| --- | --- | --- | --- | --- |
| Age at earliest overall SSI or SSDI award<sup>1</sup> |  |  | 51.79 (12.92) | 56.42 |
| Age 18-24 | 8,642 | 5.01 |  |  |
| Age 25-34 | 8,283 | 4.80 |  |  |
| Age 35-44 | 10,107 | 5.85 |  |  |
| Age 45-54 | 27,709 | 16.05 |  |  |
| Age >=55 | 105,967 | 61.38 |  |  |
| Missing | 11,932 | 6.91 |  |  |
| Time on SSI benefit roll (month)<sup>2</sup> |  |  | 81.22 (35.30) | 74.94 |
| Time on SSDI benefit roll (month)<sup>3</sup> |  |  | 76.97 (25.47) | 72.90 |
| Sex |  |  |  |  |
| Female | 81,644 | 47.29 |  |  |
| Male | 90,996 | 52.71 |  |  |
| Race and ethnicity |  |  |  |  |
| Hispanic | 16,708 | 9.68 |  |  |
| Non-Hispanic Black | 33,165 | 19.21 |  |  |
| Non-Hispanic White | 103,731 | 60.09 |  |  |
| North American Indian/Alaskan Native | 1,279 | 0.74 |  |  |
| Asian American, Pacific Islander | 3,788 | 2.19 |  |  |
| Other | 1,789 | 1.04 |  |  |
| Missing | 12,180 | 7.05 |  |  |
| Education |  |  |  |  |
| Less than high school or GED | 6,833 | 3.96 |  |  |
| High school/GED/completion cert. | 114,358 | 66.24 |  |  |
| Some college or above | 49,746 | 28.81 |  |  |
| Missing | 1,703 | 0.99 |  |  |
| Live in own house, alone, no dependents |  |  |  |  |
| Yes | 128,858 | 74.64 |  |  |
| No | 42,156 | 24.42 |  |  |
| Missing | 1,626 | 0.94 |  |  |
| Born in U.S. |  |  |  |  |
| Yes | 151,758 | 87.90 |  |  |
| No | 20,882 | 12.10 |  |  |
| Could understand, read, write English |  |  |  |  |
| Yes | 163,030 | 94.43 |  |  |
| No | 9,580 | 5.55 |  |  |
| Missing | 30 | 0.02 |  |  |
| U.S. Region |  |  |  |  |
| Northeast | 35,122 | 20.34 |  |  |
| Midwest | 36,452 | 21.12 |  |  |
| South | 63,937 | 37.04 | ☐ | ☐ |
| West | 34,857 | 20.19 | ☐ | ☐ |
| US Territories | 2,181 | 1.26 |  |  |
| Missing | 91 | 0.05 | ☐ | ☐ |
| Age at earliest award | ☐ | ☐ | ☐ | ☐ |
| Age<18 | 4,288 | 2.48 | ☐ | ☐ |
| Age≥18 | 168,352 | 97.52 | ☐ | ☐ |
| Top 5 primary diagnosis categories | ☐ | ☐ | ☐ | ☐ |
| Musculoskeletal disorders | 77,761 | 45.04 | ☐ | ☐ |
| Mental disorders | 50,244 | 29.10 | ☐ | ☐ |
| Neurological disorders | 16,951 | 9.82 | ☐ | ☐ |
| Cardiovascular system | 6,866 | 3.98 |  |  |
| Immune system disorders | 6,143 | 3.56 | ☐ | ☐ |
| Top 5 primary diagnoses |  |  |  |  |
| Disorder of the skeletal spine | 33,174 | 19.22 |  |  |
| Other and unspecified arthropathies | 21,628 | 12.53 |  |  |
| Depression, bipolar, & related disorders | 19,965 | 11.56 |  |  |
| Osteoarthritis & allied disorders | 16,333 | 9.46 |  |  |
| Neurocognitive disorder | 7,375 | 4.27 |  |  |
| Top 5 occupation groups |  |  |  |  |
| Transportation & material moving occupations | 22,523 | 13.05 |  |  |
| Production occupations | 18,906 | 10.95 |  |  |
| Office & administrative support occupations | 17,661 | 10.23 |  |  |
| Sales & related occupations | 16,492 | 9.55 |  |  |
| Food preparation, serving related occupations | 15,968 | 9.25 |  |  |
| Missing | 14,503 | 8.40 |  |  |
| Job status in the past 15 years at application |  |  |  |  |
| Always worked part-time | 18,083 | 10.47 |  |  |
| Always worked full-time | 115,920 | 67.15 |  |  |
| Worked part-time or full time | 38,637 | 22.38 |  |  |
| Total no. of jobs worked |  |  | 2.23 (1.41) | 1.00 |
| Stopped working due to health conditions |  |  |  |  |
| Yes | 119,914 | 69.46 |  |  |
| No | 52,726 | 30.54 |  |  |
| BMI categories | ☐ | ☐ | ☐ | ☐ |
| Underweight (BMI<18.5) | 3,531 | 2.05 | ☐ | ☐ |
| Healthy weight (BMI 18.5-24.9) | 40,133 | 23.24 | ☐ | ☐ |
| Overweight (BMI 25-29.9) | 52,857 | 30.62 | ☐ | ☐ |
| Class 1 obesity (BMI 30-34.9) | 38,498 | 22.30 | ☐ | ☐ |
| Class 2 obesity (BMI 35-39.9) | 19,842 | 11.49 |  |  |
| Class 3 (severe) obesity (BMI≥40) | 17,554 | 10.17 | | |
| Missing | 225 | 0.13 |  |  |
| Conditions cause pain/other symptoms |  |  |  |  |
| Yes | 158,561 | 91.85 |  |  |
| No | 13,871 | 8.03 |  |  |
| Missing | 208 | 0.12 |  |  |
| Ticket ever in use |  |  |  |  |
| Yes | 8,828 | 5.12 |  |  |
| No | 159,163 | 92.19 |  |  |
| Missing | 4,649 | 2.69 |  |  |
| Benefits ever suspended or terminated for work ☐ | ☐ | ☐ | ☐ | ☐ |
| Yes ☐ | 19,341 | 11.20 | ☐ | ☐ |
| No ☐ | 153,287 | 88.79 | ☐ | ☐ |
| Missing ☐ | 12 | 0.01 | ☐ | ☐ |
| Total amount of combined (SSDI/SSI) estimated benefits forgone for work (\$)<sup>4</sup> | | | 1,233.50<br>(7,745.79) | 0.00 |
| Total months of combined (SSDI/SSI) suspense or termination for work<sup>5</sup> |  |  | 0.77 (4.91) | 0.00 |

<sup>1</sup>n=168,263; <sup>2</sup>n=85,622; <sup>3</sup>n=129,730; <sup>4</sup>n=172,628; <sup>5</sup>n=172,611

On average, beneficiaries had worked two jobs throughout their work history. The five most common occupations were in transportation and material moving (13.05%); production (10.95%); office and administrative support (10.23%); sales (9.55%); and food preparation and serving (9.25%). Approximately two-thirds always worked full-time (67.15%), followed by those who worked both part-time and full-time (22.38%), and those who had consistently worked part-time (10.47%).

Over 5% of beneficiaries had an assigned Ticket (5.37%) and approximately 95% of that group used the Ticket at some point (5.12% of the total sample). Additionally, 11.20% of beneficiaries had their benefits suspended or terminated due to work. On average, benefits were suspended or terminated for about a month (mean=0.77, SD=4.91).

Regarding beneficiaries’ functional capacity, 47.12% had only a PRFC assessment, 42.78% had only an MRFC assessment, and 10.10% had both PRFC and MRFC assessments. Table 2 presents the results of the PRFC assessment for 98,799 beneficiaries. At the exertional level, a large proportion of beneficiaries were capable of lifting or carrying light items (70.34%); standing or walking for at least six hours without using a handheld assistive device (48.42%); and sitting for at least six hours without needing to alternate sitting and standing to relieve pain or discomfort (92.88%). However, slightly over half of the beneficiaries had limitations in performing pushing or pulling activities (51.47%), and experienced postural (89.12%), manipulative (66.58%), and environmental (61.85%) restrictions.

**Table 2.** Physical Residual Functional Capacity of Study Sample (n=98,799)

| Variable□ | Frequency□ | Percent□ | Average (SD)□ | Median□ |
| --- | --- | --- | --- | --- |
| Exertional limitations: lift & carry |  |  | N/A | N/A |
| Sedentary | 24,139 | 24.43 |  |  |
| Light | 69,495 | 70.34 |  |  |
| Medium | 862 | 0.87 |  |  |
| Heavy | 3,010 | 3.05 |  |  |
| Very heavy | 10 | 0.01 |  |  |
| Missing | 1,283 | 1.30 |  |  |
| Exertional limitations: standing/walking with/out handheld assistive device (AD) use |  |  | N/A | N/A |
| Stand/walk < 6 hours with AD use | 7,106 | 7.19 |  |  |
| Stand/walk < 6 hours without AD use | 41,786 | 42.29 |  |  |
| Stand/walk >= 6 hours with AD use | 785 | 0.79 |  |  |
| Stand/walk >= 6 hours without AD use | 47,840 | 48.42 |  |  |
| Missing | 1,282 | 1.30 |  |  |
| Exertional limitations: siting/standing |  |  | N/A | N/A |
| Sit < 6 hours & require alternate sitting & standing | 653 | 0.66 |  |  |
| Sit < 6 hours & not require alternate sitting & standing | 1542 | 1.56 |  |  |
| Sit >= 6 hours & require alternate sitting & standing | 3,557 | 3.60 |  |  |
| Sit >= 6 hours & not require alternate sitting & standing | 91,765 | 92.88 |  |  |
| Missing | 1,282 | 1.30 |  |  |
| Exertional limitations: pushing/pulling |  |  | N/A | N/A |
| Unlimited | 46,677 | 47.24 |  |  |
| Limited | 50,854 | 51.47 |  |  |
| Missing | 1,268 | 1.28 |  |  |
| No. of postural limitations with occasionally or never response |  |  | 5.13 (2.23)<sup>1</sup> | 6 |
| 0 | 2,130 | 2.16 |  |  |
| 1 - 7 | 7,348 | 89.12 |  |  |
| Missing | 8,611 | 8.72 |  |  |
| No. of manipulative limitations: |  |  | 1.70 (0.87)<sup>2</sup> | 1 |
| 0 | 53 | 0.05 |  |  |
| 1 - 4 | 65,777 | 66.58 |  |  |
| Missing | 32,969 | 33.37 |  |  |
| No. of visual limitations |  |  | 2.93 (1.79)<sup>3</sup> | 2 |
| 0 | 4 | 0.00 |  |  |
| 1 - 6 | 7,769 | 7.87 |  |  |
| Missing | 91,026 | 92.13 | 1.04 (0.21)<sup>4</sup> | 1 |
| No. of communicative limitations |  |  |  |  |
| 0 | 0 | 0.00 |  |  |
| 1 - 2 | 3,590 | 3.63 |  |  |
| Missing | 95,209 | 96.37 |  |  |
| No. of concentrated, moderate, or all environmental limitations |  |  | 2.55 (1.75)<sup>5</sup> | 2 |
| 0 | 0 | 0.00 |  |  |
| 1 - 8 | 61,108 | 61.85 |  |  |
| Missing | 37,691 | 38.15 |  |  |
| <sup>1</sup>n= 90,188; <sup>2</sup>n= 65,830; <sup>3</sup>n= 7,773; <sup>4</sup>n= 3,590; <sup>5</sup>n= 61,108 |  |  |  |  |

Of the 91,284 beneficiaries who had an MRFC assessment in 2016, more than half had no significant limitations in any of the four assessment categories (Figure 1). However, a considerable proportion of beneficiaries had serious limitations in sustained concentration and persistence (45.54%), followed by adaptation (20.38%), understanding and memory (19.89%), and social interaction (18.87%). Notably, the last three categories had over 25% missing data, either because the category had not yet been answered or was unratable due to insufficient evidence.

**Figure 1.**
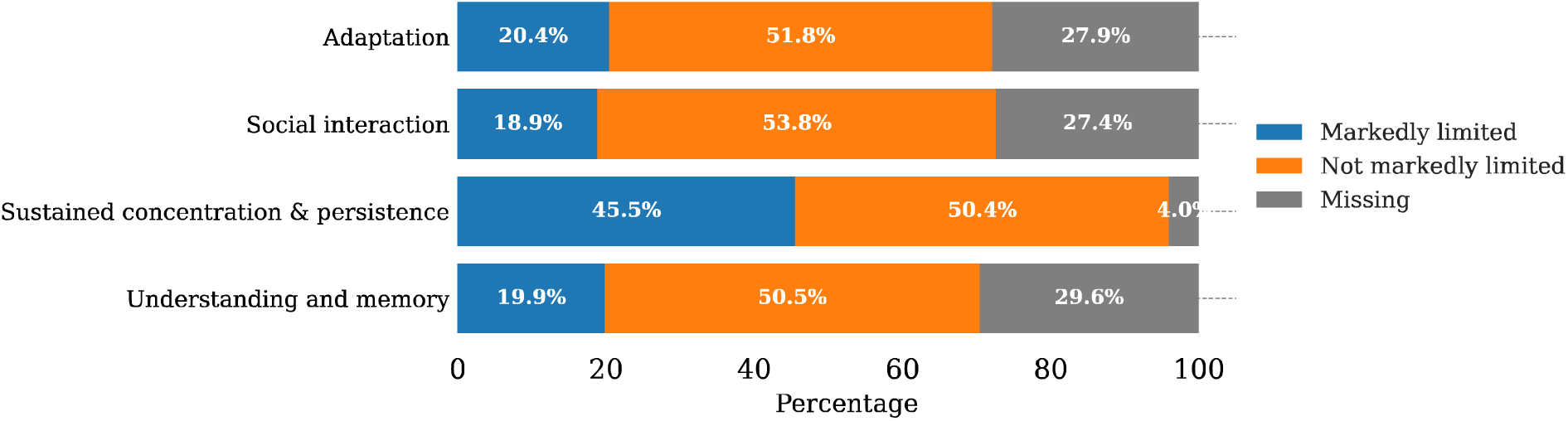
Distribution of functional limitations in each of four categories of mental function.

### TTW program participation

In both frequentist and Bayesian analyses, we used logistic regression to estimate the odds of Ticket participation among beneficiaries, categorized by SSA program and RFC assessment type. A contingency table for these two grouped variables is available in the Supplemental Materials. Figure 2 and Figure 3 present the odds ratios for the top eighteen most impactful predictors in the frequentist and Bayesian models, respectively.

**Figure 2.**
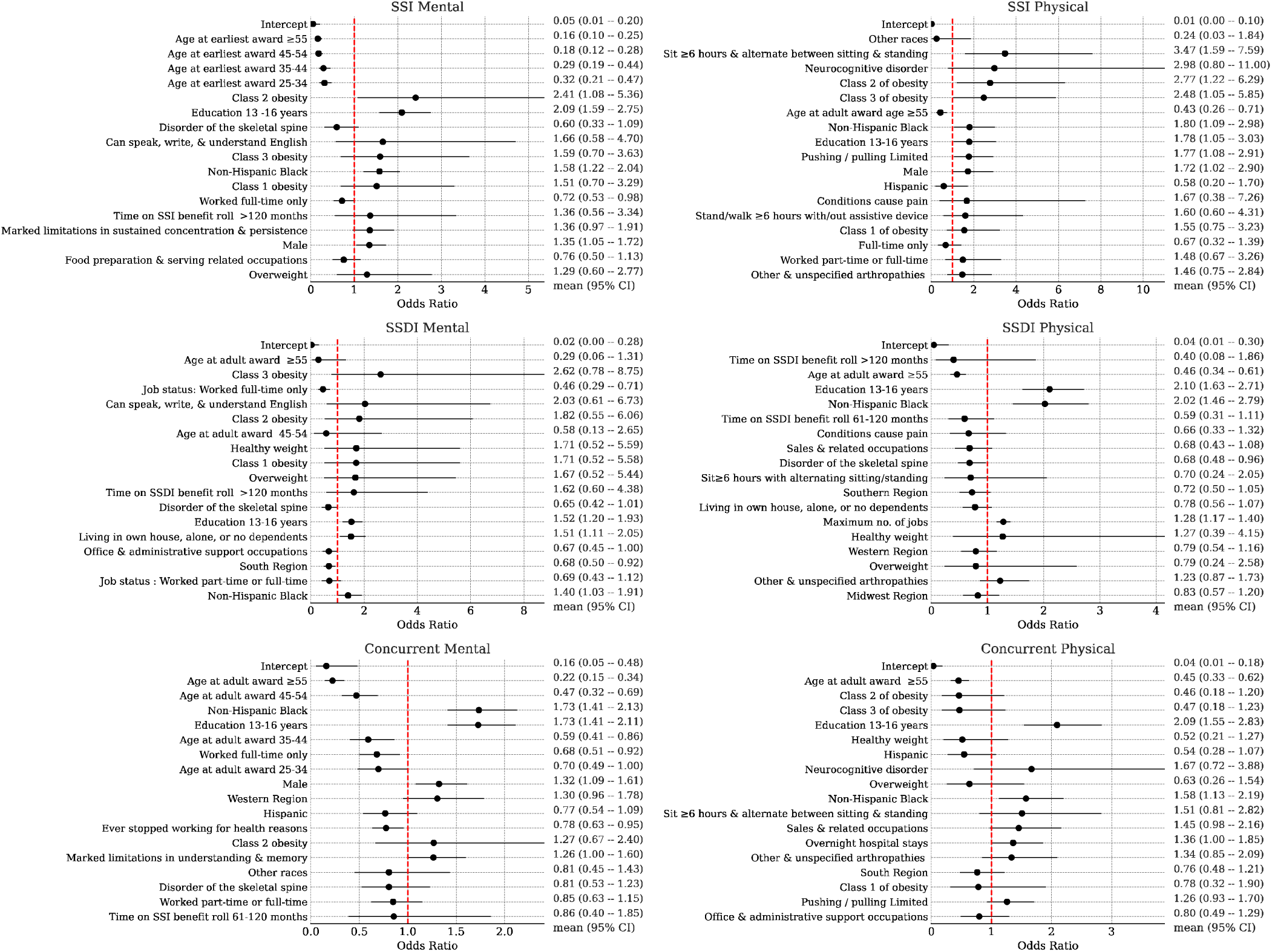
Frequentist logistic regression model for the instrument of ticket utilization. Shown are odds ratios and 95% confidence intervals for the top eighteen most-impactful predictors (ranked by absolute log odds ratios) by model.

**Figure 3.**
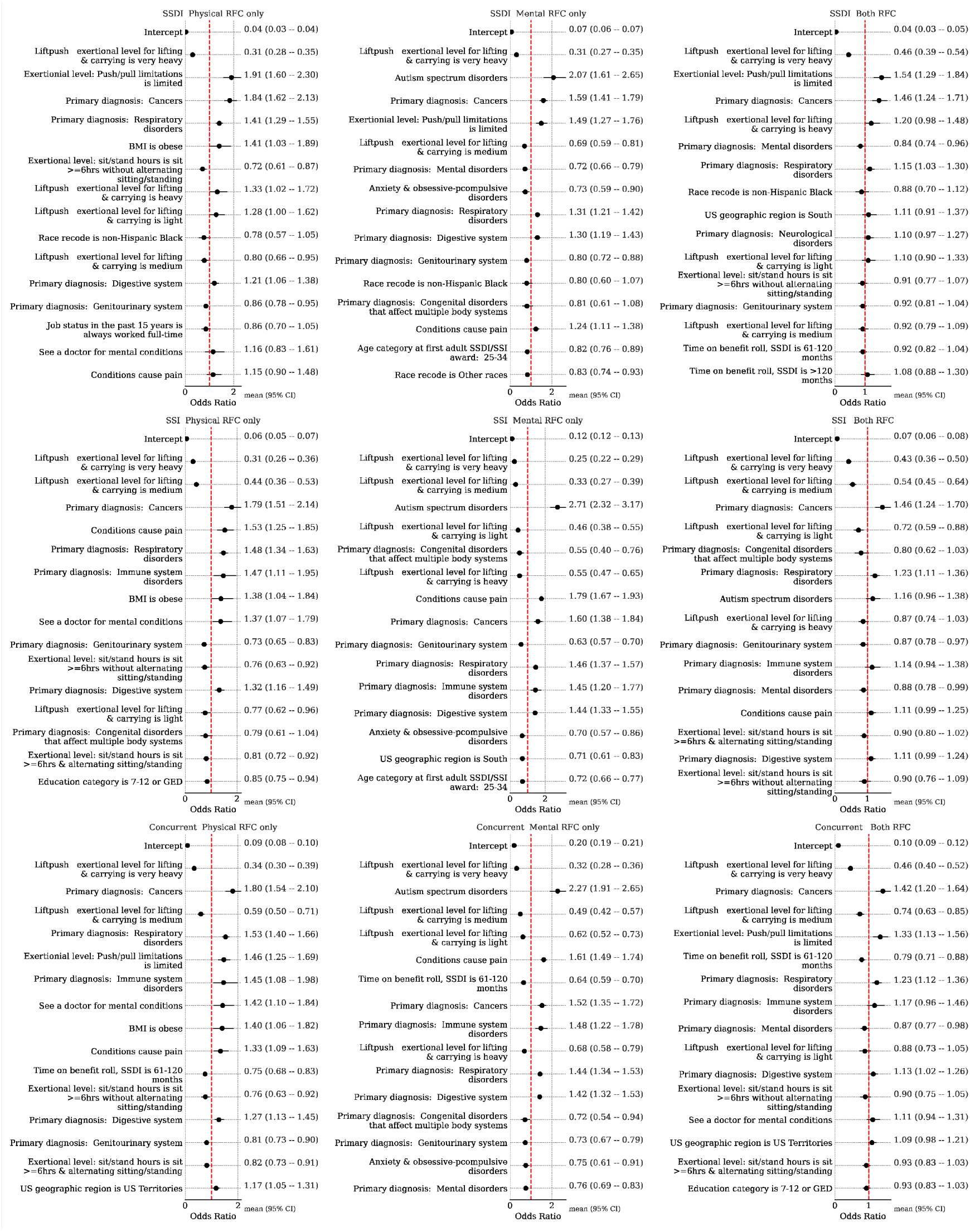
Bayesian logistic regression model coefficients (exponentiated into odds ratios) for ticket participation prediction. Mean and middle 95% credible region are shown.

In the frequentist analyses, we fit separate models based on RFC availability, as not all beneficiaries had both types of assessments. These analyses identified consistent characteristics among beneficiaries more likely to participate in the TTW program, regardless of program type or RFC. Key predictors included being non-Hispanic Black and having a college of education. Additionally, a history of working multiple jobs was associated with increased odds of participation, except among SSI beneficiaries with a PRFC assessment. Among SSI beneficiaries, being male and obese were also linked to higher participation. In contrast, older age at the time of receiving initial disability benefits was associated with decreased odds of participation across all groups, except SSDI beneficiaries with an MRFC assessment. Other negative predictors of TTW participation included living in the South, ceasing work due to health-related issues, and a history of full-time employment. Health and functional conditions, such as skeletal spine-related disorders, limitations in social interaction, and environmental limitations, were also linked to lower odds of participation.

The Bayesian analyses largely confirmed these socio-demographic characteristics as predictors of participation. In addition, they highlighted other factors associated with a higher likelihood of TTW participation, including residing in U.S. territories and conditions like obesity, pain, cancers, respiratory disorders, autism, and mental health-related doctor visits. Exertional limitations, such as heavy lifting/carrying capacity, were generally associated with lower participation.

### Benefit suspension or termination due to work

Figure 4 presents the top factors predicting work-related benefit suspension, ranked by odds ratios (ORs) and 95% confidence intervals, based on the frequentist models, broken down by cohort. Figure 5a presents these factors according to the Bayesian model, while Figure 5b presents the doubly robust estimates for the effect of TTW participation on the primary outcome. Both analytic approaches consistently revealed a clear impact of TTW participation on the likelihood of benefit suspension or termination, with the Bayesian model estimating larger treatment effects compared to the frequentist model.

**Figure 4.**
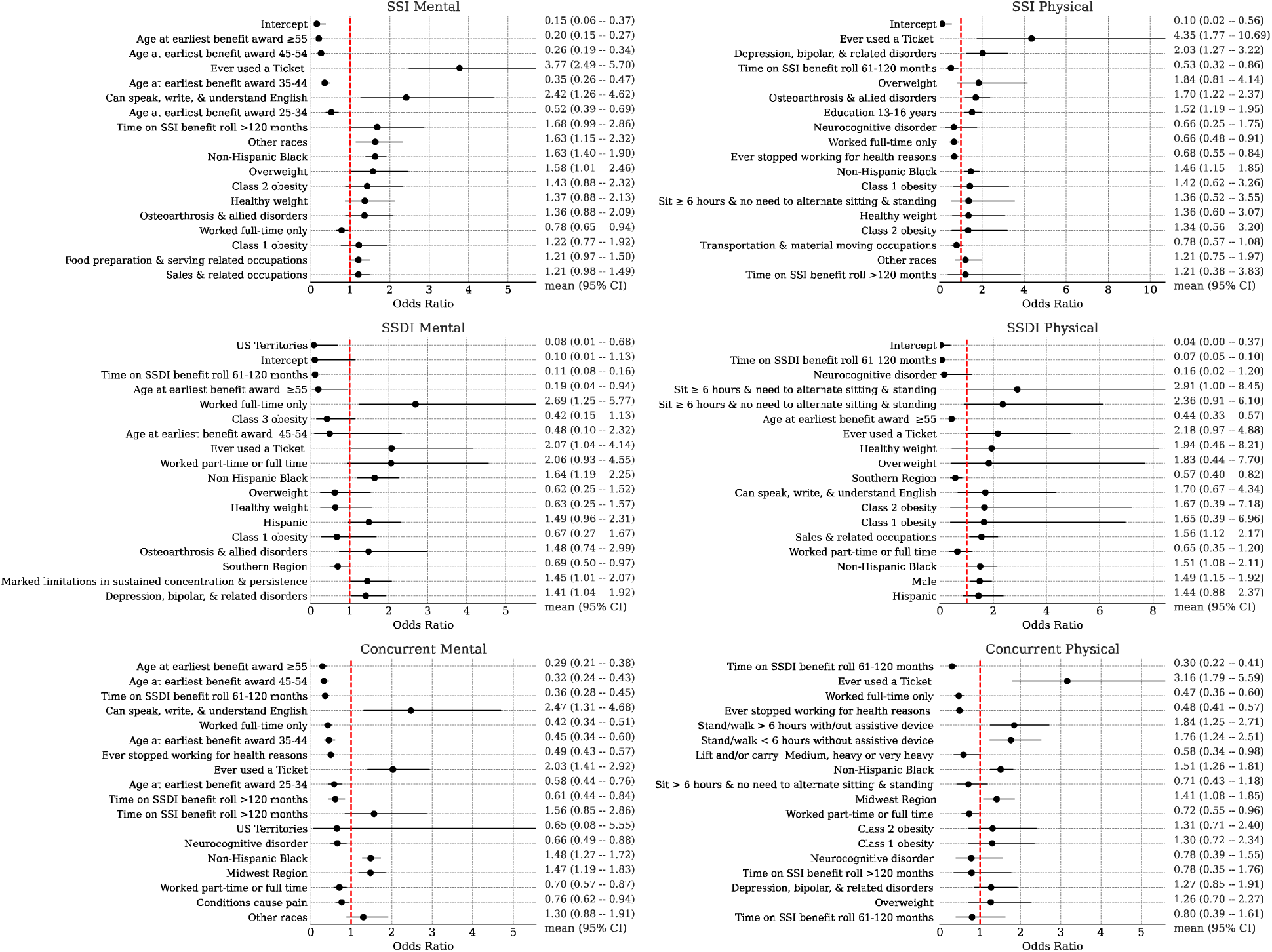
Frequentist logistic regression model for the primary outcome of benefits cessation due to work. Shown are odds ratios and 95% confidence intervals for the top eighteen most-impactful predictors (ranked by absolute log odds ratios) by model.

**Figure 5.**
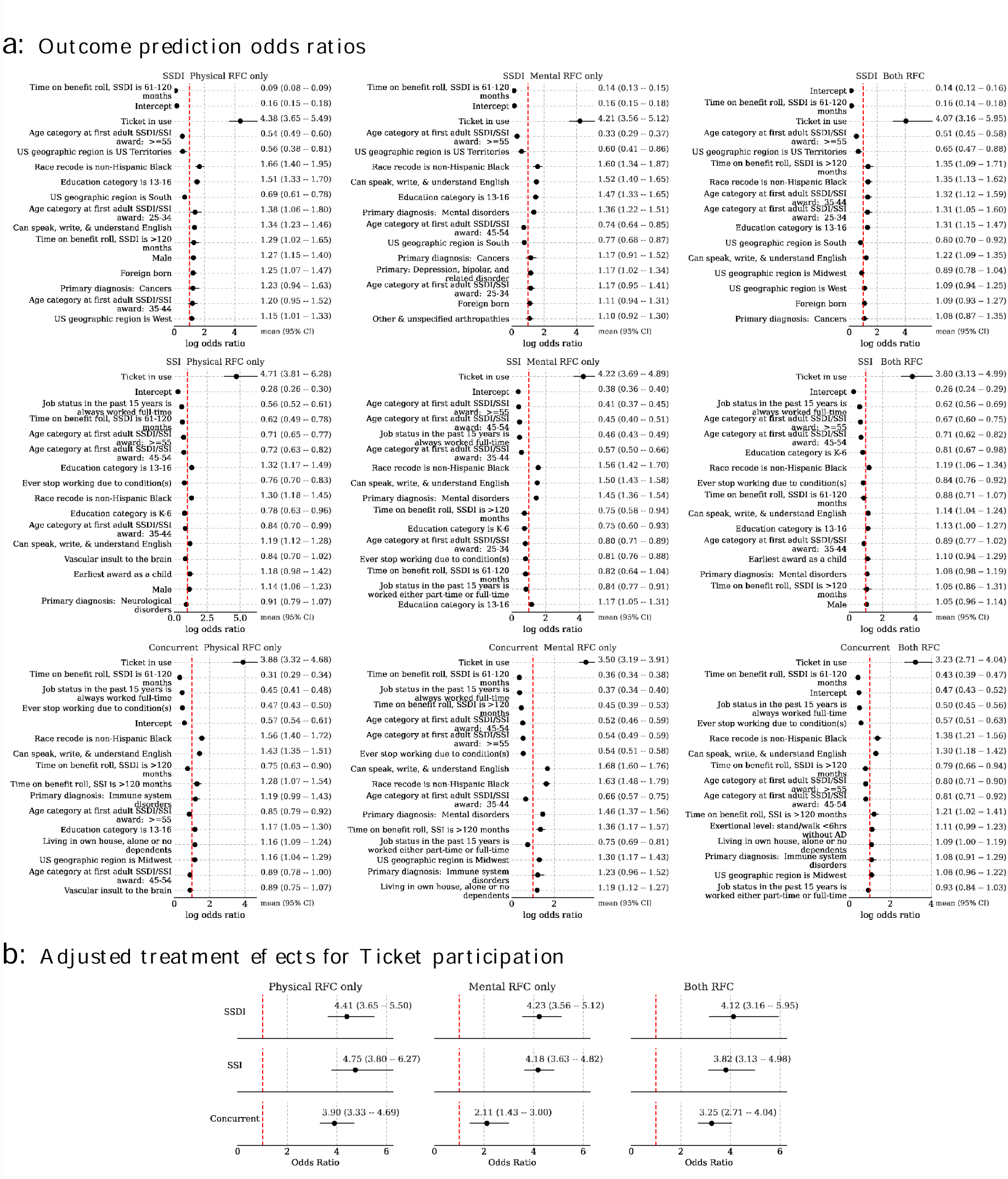
a) Bayesian logistic regression model coefficients (exponentiated into odds ratios) for predicting benefits cessation due to work. b) Bayesian doubly robust estimates of Ticket to Work average treatment effects conditional on SSI/SSDI status and Physical/Mental RFC presence. Mean and middle 95% credible region is given.

The frequentist analysis indicated that among 6,172 matched SSI beneficiaries with an MRFC assessment, Ticket users had notably higher odds than non-users of experiencing a work-related benefit suspension or termination (OR=4.90; 95% CI:3.43-6.98). A similar trend was observed among 3,481 matched SSI beneficiaries with a PRFC assessment (OR=5.38, 95% CI: 2.46-11.80). In line with these results, the Bayesian models estimated an OR of 4.18 (95% CrI: 3.63-4.82) for MRFC and 4.75 (95% CrI: 3.80-6.27) for PRFC. For beneficiaries with both MRFC and PRFC, the estimated OR was 3.82 (95% CrI:3.13-4.98).

For 8,763 matched SSDI beneficiaries with an MRFC assessment, the frequentist model showed that Ticket users had 3.51 times higher odds of a work-related benefit suspension or termination than non-users (OR=3.51, 95% CI=1.92-6.42). A similar pattern was observed in 14,274 matched SSDI beneficiaries with a PRFC assessment (OR=3.13, 95% CI=1.65-5.93). Bayesian models produced consistent results, with estimated ORs of 4.23 (95% CrI: 3.56-5.12) for MRFC, 4.41 (95% CrI:3.65-5.50) for PRFC, and 4.12 (95% CrI: 3.16-5.95) for those with both MRFC and PRFC assessments.

Among 6,725 matched Concurrent beneficiaries with an MRFC assessment, the frequentist model revealed that Ticket users also had higher odds of work-related benefit suspension or termination than non-users (OR=2.12, 95% CI=1.61-2.78). This trend persisted among 5,312 matched Concurrent beneficiaries with a PRFC assessment (OR=2.65, 95% CI=1.62-4.34). Similar findings emerged from the Bayesian analyses, with an estimated OR of 2.11 (95% CrI:1.43-3.00) for MRFC, 3.90 (95% CrI:3.33-4.69) for PRFC, and 3.25 (95% CrI:2.71-4.04) for those with both MRFC and PRFC assessments.

## Discussion

Overall, this study demonstrates that the TTW program, as currently administered, is highly effective. The application of propensity score matching and Bayesian models provide valuable insights into the characteristics of beneficiaries who are more or less likely to participate in the program, guiding potential expansion of outreach efforts. Despite the lack of data on specific vocational services or supports received by beneficiaries, both analytic approaches yielded largely consistent results that support our hypothesis. These analyses also highlighted key factors driving TTW participation and further demonstrated the program’s effectiveness in facilitating benefit suspension or termination due to work.

### TTW program participation

The frequentist analyses identified several common characteristics among beneficiaries across different SSA programs that contribute to TTW participation. These included younger age at the time of award, being non-Hispanic Black, having a college education, and a history of working multiple jobs. These findings – particularly related to age, race, and education – are consistent with previous evaluations of TTW participation. Similarly, the Bayesian analyses reinforced the importance of younger age and college education as contributing factors.

Functional characteristics also played a role in TTW participation. For example, PSM analyses revealed that SSI beneficiaries who could sit for at least six hours but require alternative sitting and standing arrangements, or those with difficulties in pushing and pulling, were more likely to participate in TTW. Among concurrent beneficiaries, individuals with serious limitations in understanding and memory were more likely to participate. The Bayesian analyses identified similar functional characteristics – such as limitations with pushing and pulling, the need for alternative sitting arrangements, and the use of assistive devices for standing or walking – as factors linked to a higher likelihood of TTW participation. While these findings may seem counterintuitive, they suggest that the TTW program may be particularly beneficial for individuals who require individually tailored workplace accommodations.

We also observed that certain health and functional characteristics negatively affected TTW participation. The PSM analyses identified skeletal spine-related conditions, health issues that led to work cessation, and significant limitations in social interaction and environmental exposure (e.g., limited tolerance to extreme cold, heat, noise, humidity, hazardous environments) as negative predictors. Many of these health-related challenges are consistent with findings from previous studies.^22,47,48^ However, unlike the frequentist analyses, the Bayesian analyses identified additional characteristics, such as conditions (e.g., cancers, pain, autism, respiratory disorders), that were associated with increased probability of Ticket participation.

### Benefit cessation due to employment

Using causal inference techniques, we quantified the impact of the TTW program on benefits suspension due to work attainment among SSI/SSDI beneficiaries. Both frequentist and Bayesian analyses reached similar conclusions, with Ticket use having a particularly large effect on benefit suspension among SSI beneficiaries. Although employment development related measures (e.g., awareness of Ticket program, job acquisition, receipt of vocationally related services) were not readily available in the datasets used, the results still support our hypothesis.

The identified characteristics can inform not only TTW policy and program development but also broader efforts to support individuals with disabilities in returning to work. By targeting individuals who are most likely to benefit from employment services, programs like TTW can enhance employment outcomes, reduce long-term reliance on disability benefits, and promote financial independence. These insights have relevance for other vocational rehabilitation and workforce development initiatives aiming to improve employment outcomes for people with disabilities. Future research is needed to evaluate the effectiveness of specific support services, identify barriers to participation across different programs, and assess the role of service providers in fostering sustained employment and long-term economic well-being.

### Limitations

This study has several limitations. First, while the PSM method is widely used in observational studies to reduce selection bias and adjust for baseline differences between groups, it can only account for observed confounders. Therefore, unmeasured confounders that influence both treatment and outcome variables may introduce residual selection bias.^49^ The Bayesian methodology, closely related to inverse propensity weighting, relies on similar assumptions. Despite this limitation, the key findings are consistent across the two results.

Second, certain variables likely influencing employment outcomes, such as marital status, awareness of Ticket program, type of support (e.g., transportation, workplace accommodations), and vocational rehabilitation services (e.g., job readiness training), were not available in the SSA administrative datasets used in this study. As a result, the direct impact of these factors on beneficiaries’ benefit suspension or termination remains unclear. Additionally, we excluded visual and communicative measures from the analytic models, as over 90 percent of beneficiaries with a PRFC assessment lacked ratings in these domains. While these data limitations restrict a comprehensive assessment of factors affecting employment and benefit outcomes, the current findings still offer valuable insights into key determinants.

Third, inconsistencies in free-form text fields posed challenges in data processing. For example, date fields occasionally contained non-date information (e.g., a season or time of year), and some text entries were truncated. Beneficiaries also reported job titles and business types interchangeably. Furthermore, certain variables exhibited little variation (e.g., understanding of English) or had very few observations in some SSDI/SSI and RFC cohorts (e.g., U.S. Territories), leading to their recoding or exclusion from some PSM models.

Finally, this study results are based solely on beneficiaries who had an RFC assessment. Therefore, the findings may not apply to beneficiaries without an RFC assessment, as their characteristics might differ.

## Conclusion

Our findings indicate that certain personal characteristics, health and functional status, and environmental factors are associated with TTW participation which, in turn, influences beneficiaries’ likelihood of benefit suspension or cessation due to work. While this study did not fully examine the impact of specific support services and vocational rehabilitation, the insights gained contribute to advancing disability research and informing policy and program development aimed at improving work participation among individuals with disabilities.

Specifically, these findings help identify potential TTW participants with a high likelihood of return-to-work success and support estimates of benefit savings for SSDI/SSI programs. Overall, this study adds to the growing body of research on service accessibility and effectiveness, which are critical for optimizing TTW outcomes and enhancing the health, independence, and quality of life for people with disabilities. Future research should further investigate the effectiveness of specific support services, identify access barriers, and evaluate factors influencing sustained employment to further improve TTW outcomes.

## CRediT authorship contributions statement

**Pei-Shu Ho**: Writing – review & editing, Writing - original draft, Conceptualization, Literature review, Data curation, Formal analysis, & interpretation. **Joshua C. Chang**: Writing – review & editing, Writing - original draft, Formal analysis, interpretation, & visualization. **Rebecca Parks:** Writing – review & editing, Literature review. **Kathleen Coale**: Writing – review & editing, Literature review. **Chunxiao Zhou**: Writing – review & editing, Data curation. **Rafael Jiménez Silva**: Writing – review & editing, Data visualization. **Julia Porcino**: Writing – review & editing, Data acquisition, Project administration. **Elizabeth Marfeo**: Writing – review & editing. **Elizabeth K. Rasch**: Writing – review & editing, Supervision, Funding acquisition.

## Declaration of generative AI in scientific writing

There is no plagiarism in this paper, including in text and images produced by AI.

## Disclaimer

The opinions and conclusions expressed are solely those of the author(s) and do not represent the opinions or policy of SSA or any agency of the federal government.

## Data availability statement

The data used in this paper are not available to the public because they include identified medical information collected by the U.S. Social Security Administration for the purposes of adjudicating claims for disability benefits and, as such, are not able to be shared.

## Data Availability

This study used data from the SSA 2021 Disability Analysis File (DAF), the electronic Claims Analysis Tool (eCAT), and the electronic Disability System (eDib). Please see the data availability links for information on entering into a data exchange with the SSA.

https://www.ssa.gov/dataexchange/request_dx.html

https://catalog.data.gov/dataset/disability-analysis-file-daf-public-use-file-puf-data-collection

## Acknowledgements

The authors would like to thank Jonathan Camacho Maldonado, MD for his support during the early stages of the project.

## Appendices A. Supplementary Figures

**Supplemental Fig. A1.**
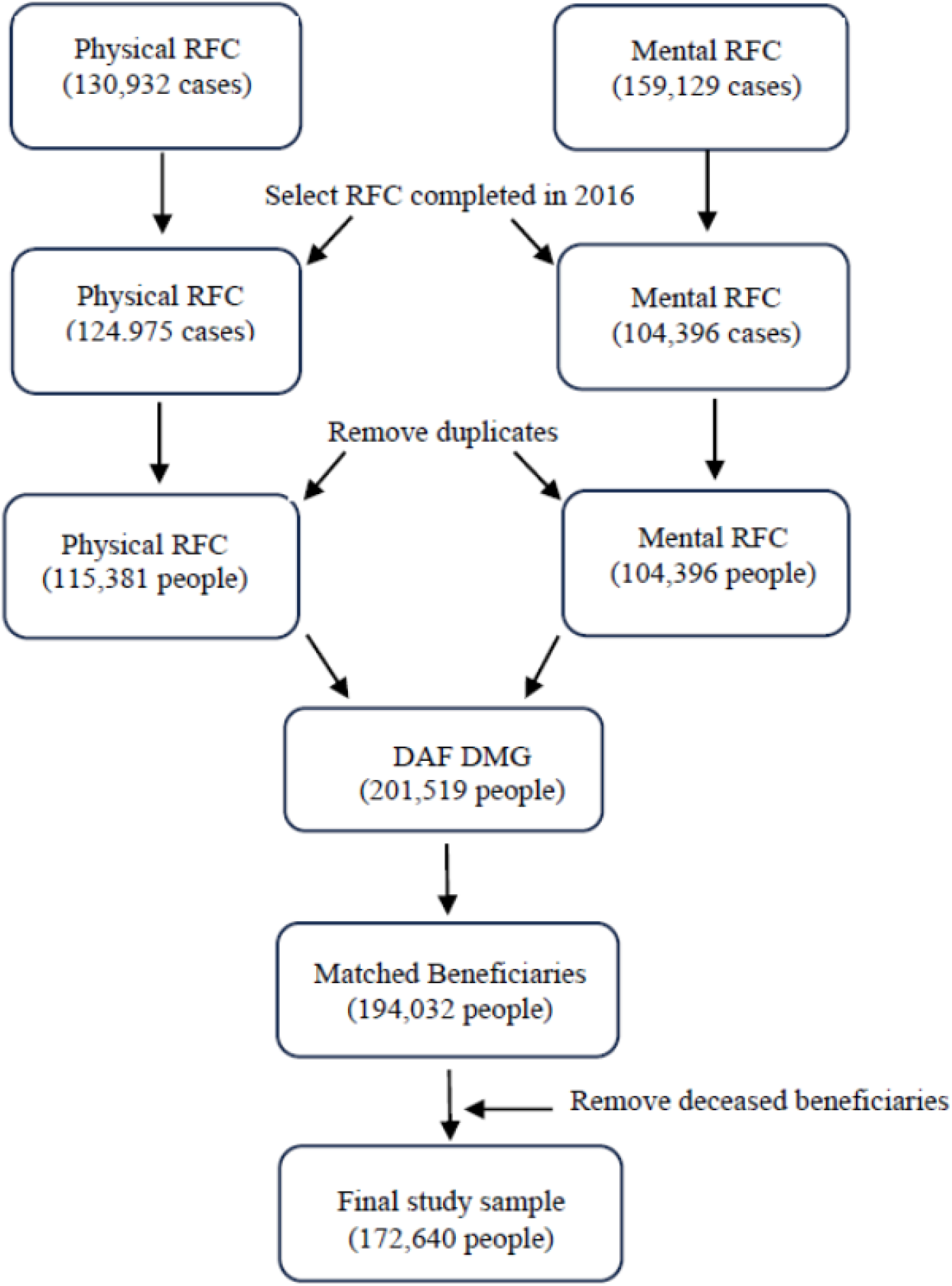
Sample Flowchart.

**Supplemental Fig. 2.**
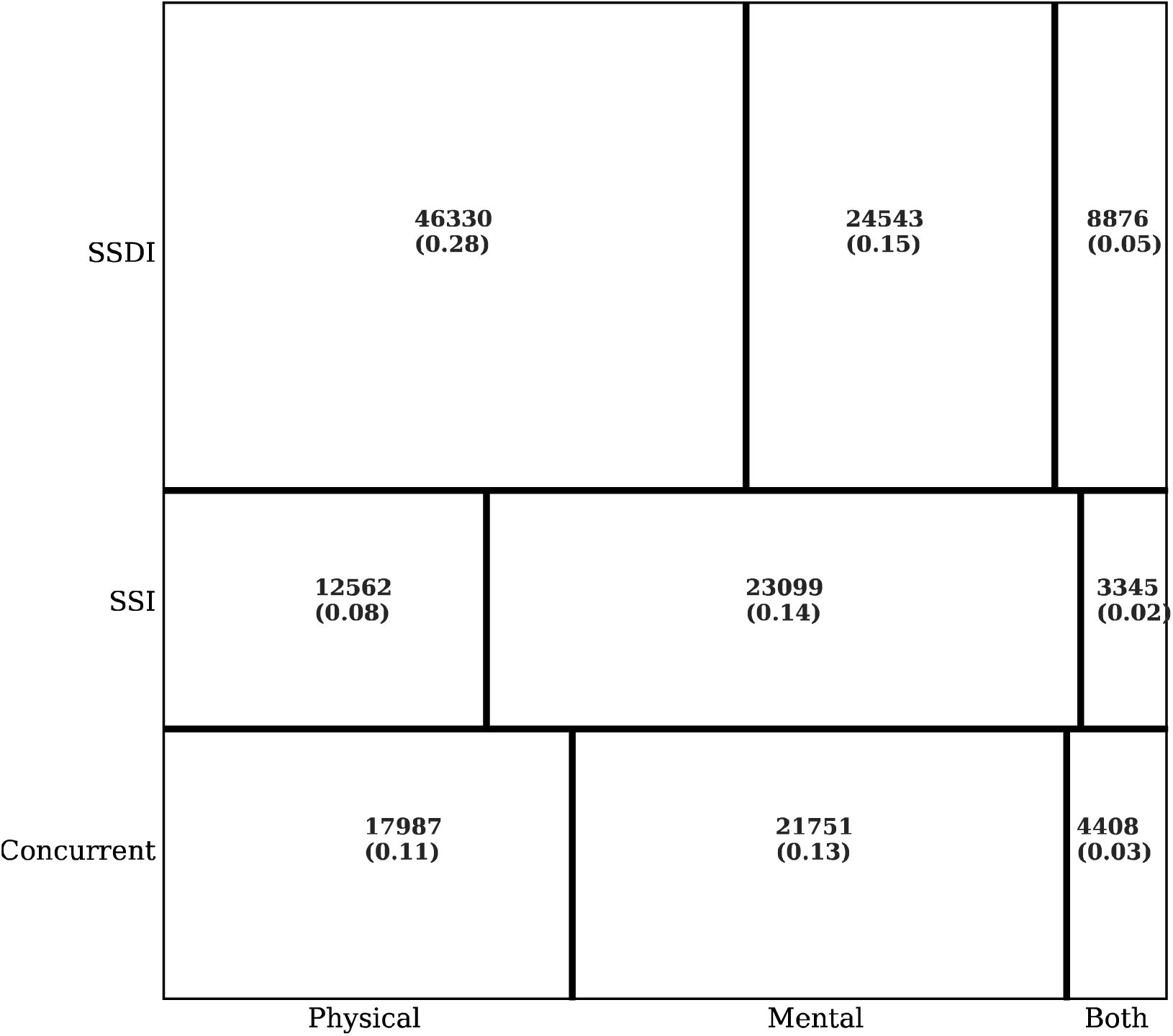
Contingency table used in parameter scaling for the Bayesian hierarchical model.

**Supplemental Fig. 3.**
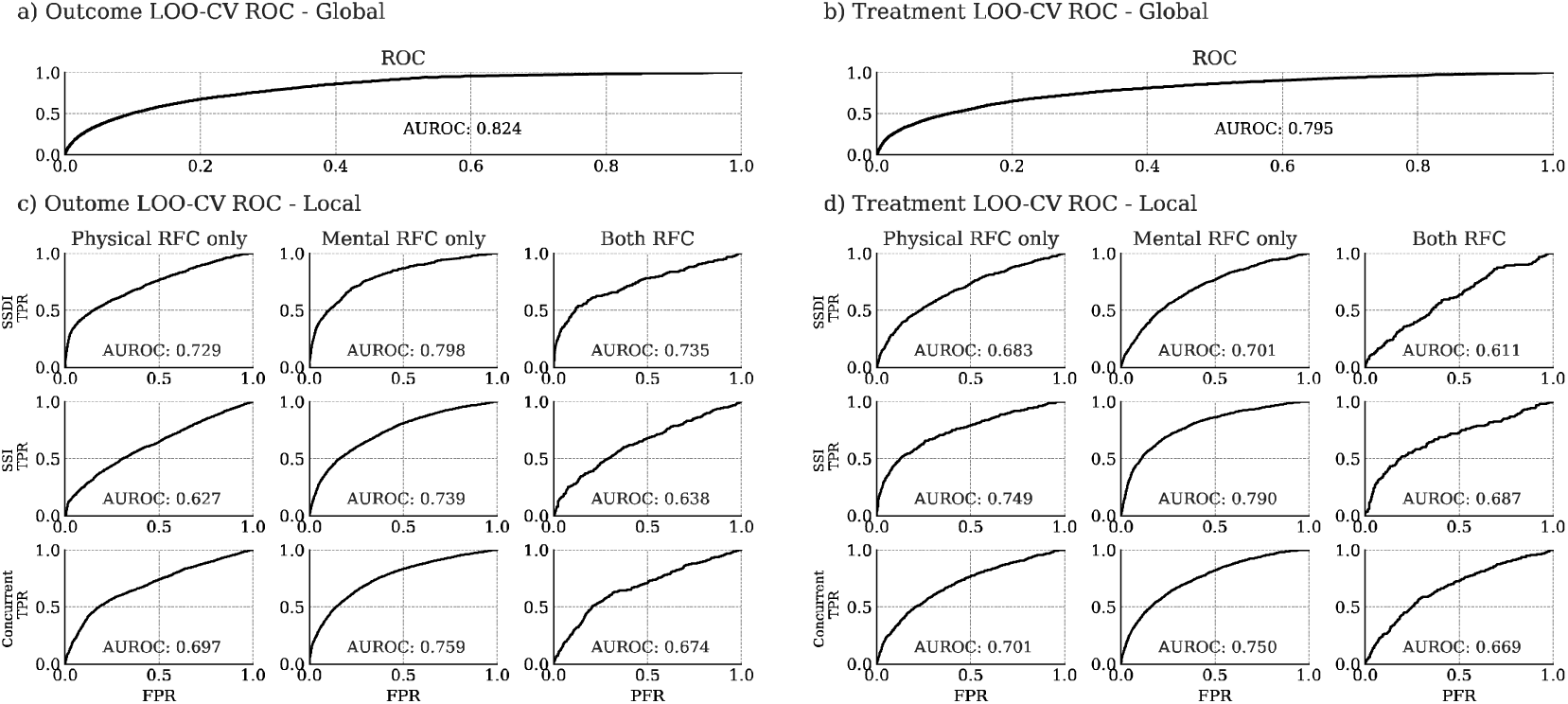
Global and local within-cohort-wise leave one out cross-validated (LOO-CV) area under the receiver (AUROC) curves for assessing the generalizability accuracy of the Bayesian joint outcome hierarchical model on both outcome and treatment assignment prediction.

## Glossary

BMI: Body Mass Index
DAF: Disability Analysis File
eCAT: Electronic Claims Analysis Tool
eDib: Electronic Disability System
EN: Employment Network
NIOCCS: NIOSH Industry and Occupation Computerized Coding System
NIOSH: National Institute for Occupational Safety and Health
PSM: Propensity Score Matching
RFC: Residual Functional Capacity
SSA: Social Security Administration
SSDI: Social Security Disability Insurance
SSI: Supplemental Security Income
SVRA: State Vocational Rehabilitation Agency
TTW: Ticket to Work
TTWIIA: Ticket to Work and Work Incentives Improvement Act

